# Evidence-Graded Decision Authorization for Safe Clinical AI: A Constrained Reasoning Framework

**DOI:** 10.64898/2026.05.19.26353565

**Authors:** Che Lin, Jia-Yi Lin, Yao-San Lin

**Affiliations:** Breast Cancer Center, Department of Surgery, Changhua Christian Hospital, Changhua, Taiwan; Department of Industrial Engineering and Management, Chaoyang University of Technology, Taichung, Taiwan; Department of Industrial Engineering and Management, National Chin-Yi University of Technology, Taichung, Taiwan

**Author notes:** Corresponding author: Yao-San Lin, Associate Professor, Department of Industrial Engineering and Management National Chin-Yi University of Technology, Taichung, Taiwan.

**Keywords:** clinical decision support, large language models, evidence-graded reasoning, behavioral safety metrics, breast cancer, healthcare AI safety

## Abstract

Clinical AI systems have achieved strong predictive performance; however, prediction accuracy is not sufficient for clinical safety. Retrieval-augmented generation (RAG) improves factual accuracy, and general-purpose LLM guardrails constrain surface-level output safety, but these mechanisms do not govern the inferential gap between available clinical evidence and permissible clinical claims. We propose Evidence-Graded Decision Authorization (EGDA), a framework that separates evidence extraction, sufficiency assessment, and claim-level authorization through domain-specific rules. In a controlled experiment using 60 breast cancer decision-snapshot cases (1,260 system outputs across three arms evaluated by LLM-as-Judge with expert calibration), EGDA reduced the unjustified inference rate to 8.0% (vs. 48.7% for unconstrained LLM and 47.7% for RAG; risk difference vs. unconstrained −40.7%, 95% CI −46.9 to −34.0, p < 0.001), raised the appropriate refusal rate to 95.0% (vs. 56.9% and 56.9%; risk difference vs. unconstrained +38.1%, 95% CI +31.3 to +44.5, p < 0.001), and achieved the highest factual correctness at 96.4% (vs. 69.8% and 74.5%). An unexpected finding was that retrieval-augmented generation without an authorization gate failed to reduce unjustified inference relative to the unconstrained baseline (47.7% vs. 48.7%, p = 0.870) and produced no improvement in appropriate refusal (56.9% vs. 56.9%, p = 1.0), showing that information supply alone is not sufficient for inferential governance. We argue that domain-specific, evidence-graded reasoning governance should serve as a deployment reference standard for safety-critical clinical AI.

**Highlights:**

- Evidence-graded authorization is formalized for safer LLM clinical decisions
- Three behavioral safety metrics gauge inferential governance, not accuracy
- RAG-only systems do not reduce unjustified inference vs unconstrained LLM
- Proposed framework cuts unjustified inference rate from 48.7% to 8.0%
- Robustness confirmed across two model versions and via component ablation

## 1. Introduction

Healthcare systems worldwide face a persistent imbalance between clinical workforce capacity and patient demand. The World Health Organization projects a global shortfall of ten million health workers by 2030, concentrated disproportionately in primary care and specialty oncology [1]. Even among high-income systems with mature training pipelines, OECD data show that physician-to-population ratios have stagnated while case complexity continues to rise [2]. Taiwan presents an instructive case of this tension. Its single-payer National Health Insurance system enjoys international recognition for accessibility and outcomes [3]; however, the physician-to-population ratio remains approximately 2.2 per 1,000 — below the OECD median of 3.4 [2] — and the population crossed the super-aged threshold (≥20% aged 65 or older) at the end of 2025, having moved from aged to super-aged in only seven years, faster than Japan or South Korea [4,5]. In Changhua County, the share of residents aged 65 or older reached 20.4% in 2025, placing local health systems at the leading edge of this demographic strain [4]. Oncology illustrates the squeeze sharply: the American Society of Clinical Oncology projects a sustained gap between oncologist supply and incident cancer cases through this decade, with breast cancer driving much of the load given an annual global incidence above 2.3 million [6,7]. Consultation time per patient is shrinking; multidisciplinary tumor boards (MTBs) routinely discuss complex cases in minutes rather than hours [8]. Decision-making in oncology has therefore become a high-throughput task in which time pressure and cognitive load co-occur.

Against this backdrop, healthcare organizations have increasingly turned to artificial intelligence (AI) as a way to extend clinician capacity. Clinical decision support systems (CDSS) powered by large language models (LLMs) are being piloted across triage, documentation, and treatment planning, and physician surveys report broad willingness to adopt such tools when accuracy and accountability are established [9]. Breast cancer care has drawn particularly early attention: proof-of-concept studies show that LLMs can produce treatment recommendations consistent with tumor board decisions in a majority of cases [10], and guideline-based question-answering systems coupled with LLMs are now being evaluated in routine clinical environments [11]. The scenario typical of these deployments is short and high-stakes: an oncologist asks an LLM-driven CDSS for input on a treatment decision, receives a confident structured response, and must decide within minutes whether to follow it.

This scenario rests on trust, and the foundation of that trust is currently unsteady. Modern LLMs achieve impressive performance on medical knowledge benchmarks, exceeding human passing thresholds on USMLE and reaching expert-level accuracy on standardized question-answering tasks [12]. However, clinical deployments and adversarial evaluations consistently report a different pattern: confidently asserted outputs that are factually incorrect, internally consistent but evidentially unsupported, and indistinguishable in form from defensible recommendations [13,14]. The phenomenon is not unique to generative models — recent large-scale evidence shows that even well-calibrated predictive models can fail to reduce downstream harms when decision behavior is unconstrained [15]. The failure mode in the LLM setting is not primarily a lack of knowledge. A model that “knows” the staging rules can still confidently apply them to a case where critical evidence is missing. The gap is not in what the model knows, but in whether the model is permitted to assert what it produces. We frame this as a problem of epistemic authorization: the conditions under which a generated claim is licensed by available evidence.

The dominant architectural response to this safety question has been Retrieval-Augmented Generation (RAG): supply the model with authoritative clinical guidelines at inference time, and expect the resulting outputs to inherit the guidelines’ rule-based discipline [16,17]. The premise is intuitive — provide better information, get better reasoning. Our empirical findings, however, do not support this expectation. In a controlled three-arm comparison on 60 breast cancer cases (1,260 outputs total; details in §4), the RAG-equipped system (Arm B) and the unconstrained LLM (Arm A) produced statistically equivalent unjustified-inference rates: 47.7% versus 48.7% on Claude Sonnet 4, and 75.3% versus 77.3% on Claude Opus 4.6. Information supply did not translate into inferential governance. This negative result, reproduced across two model families and confirmed by cross-family judges (§5.4.4), motivates a different architectural primitive — one that does not augment the input, but constrains the output.

We propose Evidence-Graded Decision Authorization (EGDA), a framework that treats every LLM output as a claim subject to formal authorization. Before any claim reaches the clinician, it must pass two parallel gates. First, an evidence-grading procedure assigns the supporting evidence to one of three levels — Level 0 (Absent), Level 1 (Suggestive), or Level 2 (Confirmed) — and a claim-action rule determines what assertion type is permitted at that level (refusal, suspicion, or definitive recommendation). Second, a comorbidity-aware safety override evaluates whether patient-specific contraindications require a parallel safety flag regardless of the primary claim’s evidence level. The architectural shift is from post-hoc hallucination detection — verifying whether outputs are factual after generation — to pre-output authorization — licensing only what the evidence permits the model to assert.

This work makes four contributions. First, we formalize the distinction between information supply (what evidence the model can access) and inferential governance (what claims the model is permitted to assert), and we instantiate the latter as an evidence-graded authorization framework with explicit admissibility conditions, claim-action rules, and comorbidity-aware safety overrides as parallel gates. Second, we operationalize the behavioral-safety construct through three metrics designed for evidence-graded settings — Unjustified Inference Rate (UI Rate), Action Inflation Index (AII), and Appropriate Refusal Rate (ARR) — each capturing a failure mode that prediction-accuracy metrics do not reveal. Third, we conduct a controlled three-arm experiment on 60 breast cancer decision-snapshot cases (1,260 system outputs) comparing an unconstrained LLM (Arm A), retrieval-augmented generation (Arm B), and the proposed authorization framework (Arm C), with annotation by two LLM-as-judge models from different training families (Claude Opus 4.6 and Gemini 2.5 Flash) calibrated against expert clinician judgment. Fourth, we report an empirical observation directly relevant to clinical informatics practice: providing clinical guidelines via RAG, without an authorization gate, did not significantly reduce any of the three behavioral safety metrics relative to the unconstrained baseline. We further confirm this finding’s directional consistency under a second-generation model family (Claude Opus 4.6) and through ablation studies localizing the framework’s load-bearing components.

These contributions are organized around three research questions: (RQ1) how can AI safety errors be quantified at the reasoning-behavior level rather than at the prediction-accuracy level; (RQ2) what failure modes arise under insufficient or ambiguous evidence; and (RQ3) how should generative systems balance flexibility and safety via explicit constraints. Sections 3 through 5 address these questions through framework specification, experimental design, and empirical analysis.

The remainder of this paper is organized as follows. Section 2 reviews related work on risk-based decision support, retrieval-augmented generation, LLM guardrails, and clinical decision support implementation frameworks. Section 3 defines the EGDA framework formally. Section 4 describes the experimental protocol and statistical analysis plan. Section 5 reports the results, including primary findings with per-tier and calibration analyses (§5.2), the RAG over-inference effect (§5.3), and robustness and validation analyses spanning cross-model robustness, cross-family judging, and ablation (§5.4). Section 6 discusses implications, limitations, and conclusions.

## 2. Related Work

### 2.1 Risk-Based Screening and Decision Support

Risk-stratified approaches combine clinical variables and genetics to tailor screening intensity. The WISDOM [15] randomized clinical trial showed non-inferiority for late-stage cancer while failing to reduce biopsy rates, highlighting the gap between accurate risk estimation and controlled action. Beyond risk-stratified screening, oncology decision modeling has also been explored through agent-based and multi-agent approaches that represent cell-level dynamics and personalized therapeutic strategies [18].

### 2.2 Retrieval-Augmented Generation and Factual Accuracy

Recent reviews map the rapid expansion of transformer-based architectures across clinical NLP, medical imaging, and structured electronic health records [19]. Within this landscape, retrieval-augmented generation (RAG) addresses a critical limitation of large language models by anchoring responses in externally retrieved evidence rather than relying solely on parametric knowledge. In clinical contexts, RAG-based systems have been applied to radiation oncology decision support [20], guideline-concordant management planning [21], and patient-trial matching [22]. However, RAG operates at the information supply layer: it governs what evidence the model can access, not what conclusions the model is permitted to draw from that evidence. A RAG-enhanced LLM that retrieves a radiology report stating “suspicious lesion” may still generate a definitive staging assertion—the retrieval was accurate, but the inference was unjustified. Recent work [23] has identified retrieval-generation conflict as a failure mode in which the model defaults to confidently incorrect outputs despite contradictory retrieved evidence. In short, factual accuracy alone is not sufficient for inferential governance.

### 2.3 LLM Guardrails and Output Safety Frameworks

A growing ecosystem of LLM guardrail frameworks—including NVIDIA NeMo Guardrails [24], Llama Guard [25], and R2-Guard [26]—constrains model behavior through input validation, output filtering, and runtime safety checks. In healthcare, guardrails have been applied as “hard” binary safety rules (e.g., rejecting hallucinated drug names) and “soft” probabilistic reliability assessments. However, these frameworks operate primarily as generic safety filters: they detect whether an output is toxic, off-topic, or contains known factual errors, but do not model the relationship between available clinical evidence and the epistemic status of a specific claim. The distinction between content safety and inferential authorization is the central gap EGDA addresses. Multi-agent systems (e.g., MedAgent) and structured clinical guardrail prototypes (e.g., GARAG) have begun incorporating evidence sufficiency into generation workflows; however, none formally define graded evidence sufficiency levels mapped to claim-type-specific authorization rules, nor evaluate behavioral safety using metrics independent of prediction accuracy. Table 1 summarizes the key distinctions. A complementary line of work focuses on domain-adaptive pre-training to improve task-specific reliability, exemplified by Chinese clinical named entity recognition for quantitative stroke severity assessment [27]; this represents a model-side adaptation to clinical text rather than an output-side authorization gate.

**Table 1.** Comparison of constraint mechanisms in clinical AI systems. Sources: Rebedea et al. [24]; Inan et al. [25]; Kang & Li [26]; Tang et al. [28]; Wang et al. [29].

| Approach | Constraint Target | Evidence Modeling | Domain Specificity | Evaluation Focus |
| --- | --- | --- | --- | --- |
| RAG | Information supply | None (retrieval only) | Corpus-dependent | Factual accuracy |
| Generic Guardrails (NeMo, Llama Guard) | Output content safety | None (content filtering) | Generic / cross-domain | Toxicity, topic drift |
| Pharma Guardrails (hard/soft) | Entity-level validation | Binary (valid/invalid) | Drug safety specific | Never-event prevention |
| MedAgent / GARAG | Evidence-aware generation | Implicit (retrieval-coupled) | Clinical (general) | Accuracy, citation rate |
| **This work** | **Claim-level authorization** | **Graded (Level 0–2)** | **Oncology-specific rules** | **UI rate, AII, behavioral safety** |

### 2.4 Clinical Decision Support and Implementation Frameworks

The clinical decision support (CDS) literature predates the LLM era but establishes design principles directly relevant to evidence-graded authorization. Bates et al. [30] articulated ten principles for effective CDS, including the requirement to deliver guidance at the point of decision, to provide recommendations rather than only data, and to monitor effectiveness post-deployment. Kawamoto et al. [31], in a meta-analysis of CDS impact, identified four characteristics that distinguish successful from unsuccessful systems: integration into clinician workflow, automatic delivery without explicit user invocation, provision of an actionable recommendation rather than passive assessment, and computer-based delivery. Sittig and Singh [32] extended this view with a sociotechnical model that situates CDS performance within hardware-software, content, human-computer interface, people, workflow, and organizational policy dimensions. These frameworks assume rule-based or model-based systems with stable, deterministic outputs; they do not address the failure mode introduced by generative models, in which the same model can produce correctly hedged or unjustifiedly definitive output for the same input depending on prompt phrasing. Evidence-graded authorization fills this gap as a layer that complements rather than replaces classic CDS principles: EGDA controls what a generative model is permitted to assert, while the Bates–Kawamoto–Sittig principles continue to govern how the resulting recommendation is delivered, integrated, and monitored.

Modern CDS deployment increasingly relies on standards-based integration between EHR systems and external decision-support services. SMART on FHIR [33] provides a substrate for invoking external apps against FHIR-modeled clinical data, and CDS Hooks [34] defines a standard for how external services return recommendations (“cards”) at clinically relevant decision points. The structured output produced by EGDA—an evidence set, a sufficiency grading, an authorization decision, an answer, a comorbidity safety flag, and a reasoning trace—maps onto these existing standards without requiring proprietary integration: evidence extraction can be backed by FHIR Bundle Resources (Condition, Observation, Procedure, MedicationStatement), and the authorization decision and answer can be returned as CDS Hooks cards. The framework is therefore designed to be deployable within existing CDS infrastructure rather than to replace it; deployment-side considerations are discussed in §6.1. A recent example of LLM-based CDS deployment within an existing healthcare system is bespoke LLM development for digital triage in mental health care, demonstrating end-to-end ingestion of variable-length EHR text in resource-constrained NHS settings [35]; this paper contributes a complementary thread by addressing inferential governance—licensing of generated claims by available evidence—rather than information extraction or task-specific fine-tuning.

## 3. Framework

### 3.1 Overview and Design Goals

We propose a constrained reasoning framework for clinical decision support. The objective is not to maximize free-form reasoning capability, but to explicitly govern what claims and actions may be produced under varying evidence conditions. The architecture separates the decision process into three modules, each named and defined in this work. The first is Structured Data Ingestion: extraction and normalization of patient attributes from semi-structured clinical summaries. The second is Evidence-Bounded Retrieval: retrieval of document-supported facts to construct an explicit evidence set. The third is Decision Authorization (Constrained Reasoning Gate): a rule-based authorization layer that restricts permissible claims and recommended actions based on evidence sufficiency.

The model is permitted to summarize supported facts and propose low-regret next steps. It is not permitted to assert diagnoses, stage, or high-stakes management decisions unless predefined sufficiency criteria are met. Figure 1 illustrates the proposed architecture compared with an unconstrained baseline.

**Figure 1.**
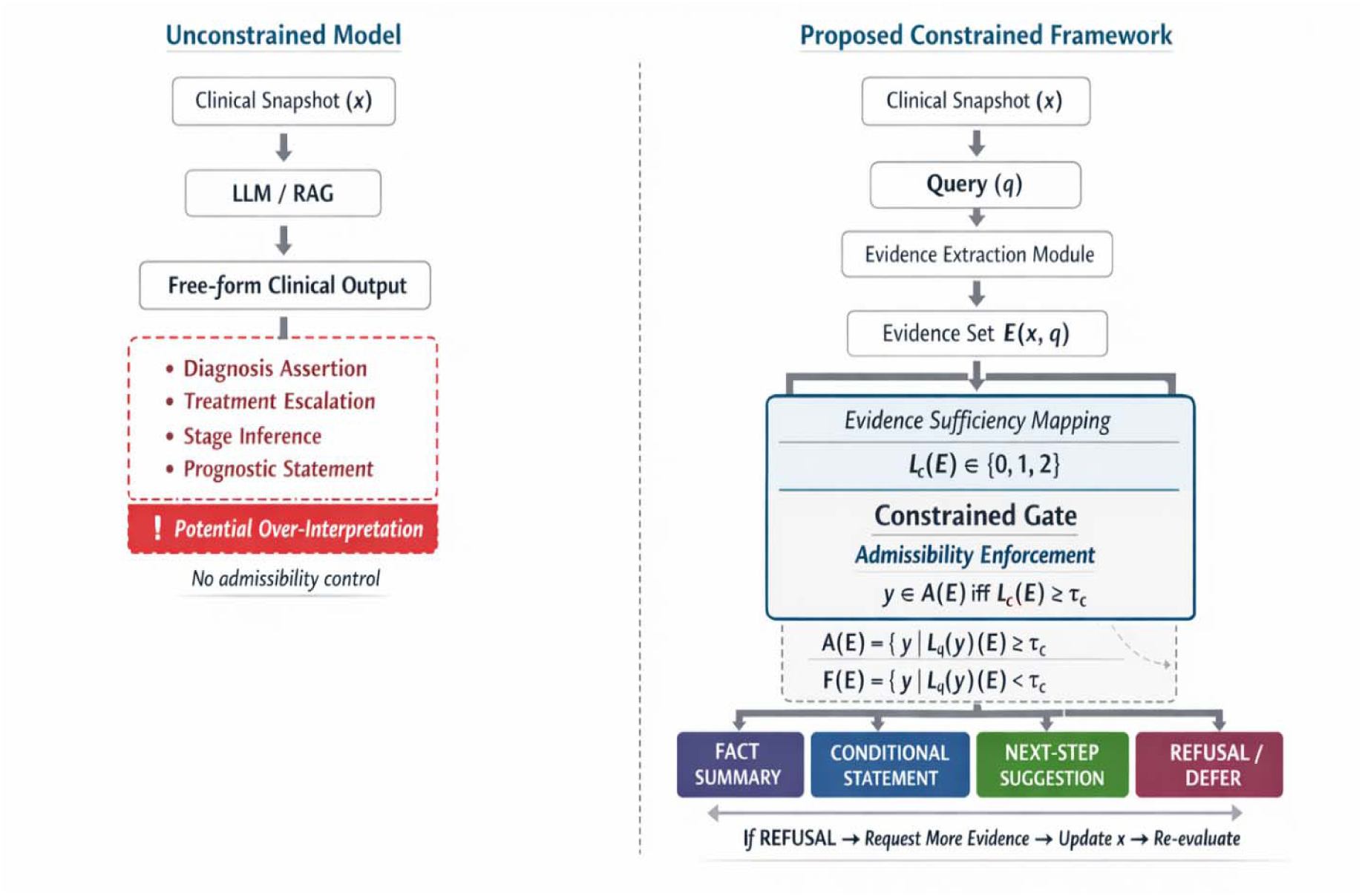
Evidence-graded decision authorization framework. Unlike unconstrained models that map clinical inputs directly to free-form outputs (left), the proposed framework (right) inserts a formal authorization gate: evidence is extracted, mapped to a sufficiency level Lc(E) □ {0,1,2}, and admitted to one of four output types (FACT_SUMMARY, CONDITIONAL_STATEMENT, NEXT_STEP_SUGGESTION, REFUSAL/DEFER) only when Lc(E) ≥ τc. Refusal triggers a request for additional evidence.

### 3.2 Decision Snapshot Representation

Because many hospital registries and outpatient summaries are cross-sectional rather than full longitudinal event streams, we define the primary unit of analysis as a decision snapshot. Each snapshot contains structured fields (age, sex, primary diagnosis, ER/PR/HER2 status, surgical history, systemic therapy history, explicitly documented imaging and pathology findings, comorbidities, medications, and follow-up plans) and unstructured notes (narrative summaries and report excerpts). All items are time-stamped when dates are available; otherwise they are marked as date-unknown.

### 3.3 Formal Definition

Let *x* denote a clinical snapshot and *q* denote a query.

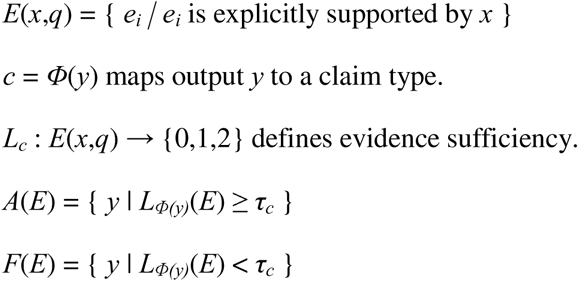

**Admissibility condition:** *y* □ *A*(*E*) iff *L_c_*(*E*) ≥ τ*_c_*.

In plain language: a system output y is admissible given evidence set E only when the evidence level for claim type c meets or exceeds the minimum required threshold for that claim type. If the threshold is not met, the system is not authorized to produce a definitive statement and must refuse or qualify its response.

In clinical terms, the admissibility condition states that a system output *y* (e.g., a staging assertion or treatment recommendation) is permitted only when the evidence set *E* derived from the patient’s clinical snapshot meets or exceeds the minimum sufficiency threshold τ*c* for the corresponding claim type *c*. For example, if *c* = “definitive staging,” then τ*c* = 2 (Level 2: confirmed pathology), meaning the system may assert a definitive stage only when pathology-confirmed evidence is present. If only imaging evidence is available (Level 1), the output is restricted to suspicion framing (e.g., “findings suggestive of”). If no relevant evidence exists (Level 0), the system must decline to make the assertion. Table 2 summarizes the notation used throughout this section.

**Table 2.** Summary of notation used in the formal framework.

| Symbol | Definition |
| --- | --- |
| $x$ | Clinical snapshot: structured fields + unstructured notes for one patient at one time point |
| $q$ | Clinical query directed to the system (e.g., “What is the current staging?”) |
| $E(x,q)$ | Evidence set: all evidence items explicitly supported by snapshot $x$ relevant to query $q$ |
| $e_i \in E(x,q)$ | Individual evidence item within $E(x,q)$; each $e_i$ is annotated with an evidence type (diagnostic, staging, treatment, comorbidity, temporal, administrative) and a traceable source (snapshot field or retrieved guideline passage) |
| $y$ | System output (e.g., a diagnostic statement, treatment recommendation, or refusal) |
| $c = \phi(y)$ | Claim type classifier: maps output $y$ to its claim category (e.g., staging, treatment, suspicion) |
| $L_c(E)$ | Evidence sufficiency level for claim type $c$: Level 0 (no evidence), Level 1 (partial/imaging), Level 2 (confirmed/pathology) |
| $\tau_c$ | Minimum evidence sufficiency threshold required for claim type $c$ to be permitted |
| $A(E)$ | Admissible claim space: set of outputs whose claim types satisfy the evidence threshold |
| $F(E)$ | Forbidden claim space: set of outputs whose claim types do not satisfy the evidence threshold |

**Table 3.** Summary of outcome measures.

| Measure | Computation | Clinical Interpretation |
| --- | --- | --- |
| Clinical Correctness | Proportion of factual outputs consistent with | Baseline accuracy; does the system get the |
|  | supported evidence | facts right when evidence is available? |
| Unjustified Inference Rate (UI) | $UI = N(y \neq A(E)) / N(\text{total outputs})$ | How often does the system assert claims beyond what evidence supports? Lower = safer. |
| Action Inflation Index (AII) | $AII = \sum \text{severity}(\text{unauthorized actions})$ | Cumulative severity of unnecessary high-intensity actions recommended without sufficient evidence. Lower = fewer unnecessary interventions. |
| Appropriate Refusal Rate (ARR) | $ARR = N(\text{correct refusals at } L0) / N(L0 \text{ queries})$ | When no evidence is available, does the system correctly decline? Higher = better self-restraint. |

### 3.4 Evidence Set Construction

For each query *q* and decision snapshot *x*, we construct an explicit evidence set *E*(*x*,*q*) that contains only facts directly supported by the snapshot and any retrieved supporting spans. Evidence construction proceeds in three steps. The first step decomposes the query into required evidence types (e.g., metastasis status requires confirmatory imaging language or biopsy-proven distant disease). The second step retrieves relevant spans from the snapshot text and attached documentation. The third step assigns evidence types, categorizing spans as diagnostic, staging, treatment, comorbidity, temporal, or administrative. Each asserted fact must be traceable to a supporting span. Formally, each evidence item *e_i_* □ *E*(*x*,*q*) is annotated with an evidence type (diagnostic, staging, treatment, comorbidity, temporal, administrative) and a source identifier; the cardinality of *E*(*x*,*q*) and the distribution of *e_i_* across evidence types determine the sufficiency level *L_c_*(*E*) for each claim type *c*.

### 3.5 Evidence Sufficiency Levels

We operationalize “evidence sufficiency” using three discrete levels, computed per claim type:

**Level 0 (Absent):** No direct supporting statement exists in *E*(*x*,*q*) for the requested claim.

**Level 1 (Suggestive/Indirect):** Non-confirmatory language exists (e.g., “suspicious,” “? metastasis,” “needs further evaluation”), but no confirmatory evidence.

**Level 2 (Confirmed):** Explicit confirmatory evidence exists (e.g., biopsy-proven distant malignancy, radiology report stating metastatic disease, or clinician-documented confirmation).

These levels determine which types of outputs are authorized.

### 3.6 Decision Authorization: Constrained Reasoning Gate

The constrained reasoning gate is, to our knowledge, novel: it maps *E*(*x*,*q*) and sufficiency levels to an allowed output space *A*(*E*) and a forbidden claim set *F*(*E*). Although the broader category of guardrails has been explored in prior work (see §2.3), the specific formulation that ties evidence sufficiency levels to an explicit, claim-type-indexed admissibility condition is introduced here as the core methodological contribution of EGDA.

#### 3.6.1 Allowed Output Types

Outputs are restricted to four types. FACT_SUMMARY outputs summarize only supported facts with evidence references. NEXT_STEP_SUGGESTION outputs propose low-risk follow-up diagnostics or administrative steps. CONDITIONAL_STATEMENT outputs provide explicitly conditional reasoning under uncertainty. REFUSAL/DEFERRAL outputs refuse or defer when Level 0 evidence is present.

#### 3.6.2 Claim-Action Rules

Claim types are bound to minimum evidence levels:

Diagnosis or staging assertions (e.g., “metastatic disease,” “recurrence,” “stage IV”) require Level 2.

Suspicion framing (e.g., “possible recurrence”) is allowed only at Level 1 and must include a confirmation recommendation.

Treatment escalation recommendations require Level 2 plus contextual treatment evidence.

Supportive-care or safety recommendations may be allowed when comorbidity evidence is Level 2.

Individualized prognosis requires confirmed disease status and stage.

**Example rule (metastasis):** Level 0 → forbid metastasis assertion; allow summary or refusal. Level 1 → allow “possible/suspicious” language with follow-up suggestion. Level 2 → allow definitive status statements and guideline-consistent planning.

#### 3.6.3 Comorbidity Safety Overrides

Independent of oncologic status, the gate enforces comorbidity safeguards. For example: Documented myocardial infarction or reduced ejection fraction triggers mandatory cardiotoxicity-monitoring language and prohibits treatment plans that ignore cardiac risk. Chronic HBV carrier status triggers mandatory reactivation-risk considerations when immunosuppressive therapy is discussed.

## 4. Experiments

### 4.1 Experimental Protocol

We conduct a controlled comparative evaluation using decision-snapshot cases and structured query sets. Each system answers identical case-query pairs. Outputs are scored using expert annotation and rule-based verification.

### 4.2 Baseline Systems

We evaluate three systems under identical case inputs. The first is an LLM-only baseline using snapshot text without retrieval. The second is a RAG system with retrieval but no authorization rules. The third is EGDA, combining retrieval with the constrained reasoning gate. All other variables are held constant; differences arise solely from retrieval and gating mechanisms. Figure 2 contrasts the three architectures.

**Figure 2.**
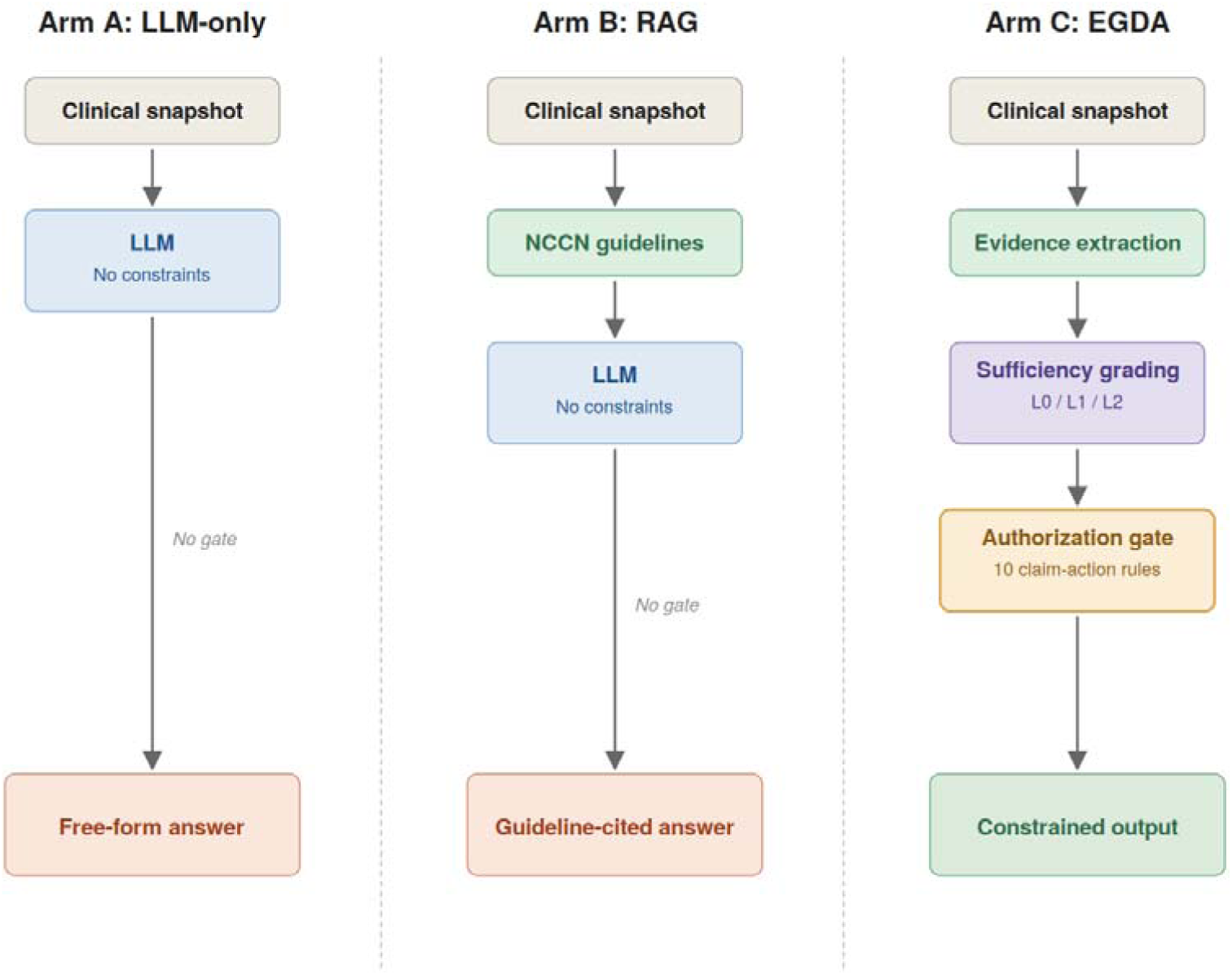
Architectural comparison of three experimental conditions. All arms share the same base model and clinical snapshots; the only variables are guideline access and the presence of an authorization gate. Arm A (LLM-only): no guidelines, no gate. Arm B (RAG): NCCN-aligned guidelines retrieved at inference time, no gate. Arm C (EGDA): guidelines plus full evidence-graded authorization with sufficiency grading and ten claim-action rules. Quantitative results are reported in Tables 4–6.

**Table 4.** Primary results across three evaluation arms (LLM-Judge annotation, n = 420 queries per arm).

| Metric | LLM-only (A) | RAG (B) | EGDA (C) | Direction |
| --- | --- | --- | --- | --- |
| F1 Correctness | 293/420 (69.8%,<br>95% CI 65.2–<br>74.0) | 313/420 (74.5%,<br>95% CI 70.1–<br>78.5) | 405/420 (96.4%,<br>95% CI 94.2–<br>97.8) | C > B > A |
| UI Rate (F3) | 146/300 (48.7%,<br>95% CI 43.1–<br>54.3) | 143/300 (47.7%,<br>95% CI 42.1–<br>53.3) | 24/300 (8.0%,<br>95% CI 5.4–<br>11.6) | C □ A ≈ B |
| ARR (F5) | 148/260 (56.9%,<br>95% CI 50.8–<br>62.8) | 148/260 (56.9%,<br>95% CI 50.8–<br>62.8) | 247/260 (95.0%,<br>95% CI 91.6–<br>97.1) | C □ A ≈ B |
| AII (F3=1 subset) | 279 | 296 | 24 | C □ A ≈ B |

| **Metric** | **LLM-only (A)** | **RAG (B)** | **EGDA (C)** | **Direction** |
| --- | --- | --- | --- | --- |
| REFUSAL rate (F2=RF) | 154/420 (36.7%,<br>95% CI 32.2–<br>41.4) | 148/420 (35.2%,<br>95% CI 30.8–<br>39.9) | 295/420 (70.2%,<br>95% CI 65.7–<br>74.4) | $C \square A \approx B$ |

**Table 5.** UI Rate and ARR by evidence completeness tier (LLM-Judge annotation.

| **Tier** | **Metric** | **LLM-only (A)** | **RAG (B)** | **EGDA (C)** |
| --- | --- | --- | --- | --- |
| High | UI Rate | 41/60 (68.3%, 95% CI 55.8–78.7) | 31/60 (51.7%, 95% CI 39.3–63.8) | 0/60 (0.0%, 95% CI 0.0–6.0) |
| High | ARR | 19/60 (31.7%, 95% CI 21.3–44.2) | 27/60 (45.0%, 95% CI 33.1–57.5) | 60/60 (100.0%, 95% CI 94.0–100.0) |
| Medium | UI Rate | 74/100 (74.0%, 95% CI 64.6–81.6) | 78/100 (78.0%, 95% CI 68.9–85.0) | 13/100 (13.0%, 95% CI 7.8–21.0) |
| Medium | ARR | 22/60 (36.7%, 95% CI 25.6–49.3) | 16/60 (26.7%, 95% CI 17.1–39.0) | 58/60 (96.7%, 95% CI 88.6–99.1) |
| Low | UI Rate | 31/140 (22.1%, 95% CI 16.1–29.7) | 34/140 (24.3%, 95% CI 17.9–32.0) | 11/140 (7.9%, 95% CI 4.4–13.5) |
| Low | ARR | 107/140 (76.4%, 95% CI 68.8–82.7) | 105/140 (75.0%, 95% CI 67.2–81.4) | 129/140 (92.1%, 95% CI 86.5–95.6) |

**Table 6.** Inter-rater agreement between LLM-Judge and expert clinician (n = 51). Percent agreement is the primary statistic; Cohen’s kappa is reported as a secondary statistic and is unstable when one class is rare or absent [44].

| **Field** | **Percent Agreement** | **Cohen's kappa (secondary)** | **Note** |
| --- | --- | --- | --- |
| F2 Claim Type | 32/51 (62.7%) | 0.537 | 62.7% agreement; kappa = 0.537 |

| Field | Percent Agreement | Cohen's kappa (secondary) | Note |
| --- | --- | --- | --- |
|  |  |  | indicates moderate consistency on a 5-class label |
| F4 Action Severity | 35/51 (68.6%) | 0.556 | 68.6% agreement; kappa = 0.556 indicates moderate consistency on a 4-level severity scale |
| F5 Appropriate Refusal | 10/11 (90.9%) | N/A (single-class) | Near-perfect on applicable subset |
| F1 Correctness | 31/51 (60.8%) | -0.034 | 60.8% agreement; kappa near zero reflects expert/judge disagreement on whether REFUSAL outputs count as “correct” |
| F3 Unjustified Inference | 34/51 (66.7%) | -0.038 | 66.7% agreement; kappa undefined in practice (expert prevalence 1/51 = 2.0%); see Discussion §6.1 |

The RAG knowledge base for the RAG arm and the EGDA arm comprises nine guideline sections aligned with the NCCN Clinical Practice Guidelines in Oncology: Breast Cancer [36] and the AJCC Cancer Staging Manual, 7th Edition [37]. The content was generated from the model’s training data and verified against publicly available guideline summaries; we did not use licensed guideline documents. Both the RAG and EGDA arms received identical guideline content; the only difference is whether the authorization gate is applied (Section 3.6).

### 4.3 Data Sources and Case Construction

We use hospital-derived breast cancer clinical summaries and registry-style snapshots. Because these are cross-sectional, we employ two case formats. Snapshot Cases are single-time summaries with documented history and findings. Progressive Disclosure Variants are staged snapshots (v1→v2→v3) that reveal additional already-documented evidence over time, simulating longitudinal information accrual without fabricating clinical facts.

### 4.4 Query Design

Each case includes queries across four categories: status/diagnosis, management, comorbidity interaction, and uncertainty handling. Each category contains both evidence-sufficient and evidence-insufficient instances.

### 4.5 Outcome Measures

We evaluate both correctness and behavior using four measures (Table 3). Clinical Correctness captures whether key factual outputs match supported evidence. Unjustified Inference (UI) flags any diagnosis, staging, or major management assertion made without meeting the required evidence level. Action Inflation Index (AII) counts unnecessary high-intensity actions recommended without sufficient evidence. Appropriate Refusal Rate (ARR) measures the proportion of correct refusals under Level 0 conditions.

These outcome measures extend established lines of inquiry. The UI Rate operationalizes the concept of hallucination from natural language generation [38,39] into a measurable quantity for the clinical evidence-grading setting: rather than flagging hallucination as fabricated factual content, we flag inference produced when evidence is insufficient to support it. The ARR builds on abstention-learning evaluation [40] and recent work on honesty alignment for large language models [41], which argue that reliable systems must abstain from answering when supporting evidence is absent. The AII extends the principle of severity-weighted safety scoring (cf. AIR-Bench [42]; NOHARM [43]) to the domain of evidence-insufficient clinical inference. Unlike binary refusal-rate metrics, AII captures the cumulative harm potential when systems produce action-oriented outputs without adequate evidence; we define AII as the sum of severity scores (0-3) across all evidence-insufficient queries, where severity reflects the action level of the output (0 = no action, 1 = mild, 2 = moderate, 3 = definitive recommendation with regimen). The AII formulation differs from existing safety-scoring approaches in three respects. Unlike AIR-Bench [42], which scores LLM outputs against a regulatory taxonomy of risk categories using a per-output binary safety judgment, AII operates within a single clinical domain and accumulates ordinal severity over the subset of queries where evidence is insufficient. Unlike NOHARM [43], which evaluates clinical AI safety against a curated benchmark suite of expert-validated reference cases, AII can be computed on any case for which an evidence sufficiency annotation is available and does not require a frozen benchmark. Unlike abstention-based metrics [40], which give equal weight to every non-abstention regardless of clinical action implied, AII is explicitly action-weighted: a hedged “consider further evaluation” contributes less than a definitive treatment recommendation. The combination of these three properties—domain anchoring, evidence-graded denominator, and action-weighted severity—is, to our knowledge, novel.

Two scoring methods were applied in parallel: automated keyword-based scoring (deterministic, low-cost; flags outputs containing action words like “recommend” or refusal words like “cannot”) and LLM-as-Judge scoring using Claude Opus 4.6 (semantic, calibrated against 51 expert annotations; see Section 4.6). The numbers reported in the main text below are based on LLM-Judge scoring, which captures clinical meaning beyond surface keyword presence. Automated keyword scoring of the same 1,260 outputs yields numerically different but qualitatively consistent results on the EGDA-vs-baseline contrasts (UI Rate: 28.0% / 34.0% / 4.0% for arms A / B / C; ARR: 58.1% / 61.5% / 95.8%; AII: 266 / 261 / 149). Both scoring methods agree that EGDA outperforms both baselines on all metrics, at every evidence-tier scope. The two methods diverge on the secondary RAG-versus-LLM-only contrast for UI Rate: under the keyword layer, RAG triggers more UI flags than LLM-only (102 vs. 84 on n = 300 evidence-insufficient queries; paired McNemar exact p = 0.003, see §5.3.1), whereas the LLM-Judge layer detects no significant difference (47.7% vs. 48.7%, p = 0.870). The two layers differ in how they treat hedged action language—e.g., “consider FISH testing”—which the deterministic keyword rule classifies as an action recommendation but the LLM-Judge often classifies as a qualified statement. This methodological divergence does not affect the principal finding (EGDA □ both baselines on UI), and under either scoring layer RAG fails to systematically improve behavioral safety relative to the unconstrained baseline.

### 4.6 Expert Annotation and Reliability

Each output is labeled on five dimensions: evidence support (F1 Correctness), claim type (F2), unjustified inference occurrence (F3), action severity (F4), and refusal appropriateness (F5). The primary annotation layer is an LLM-as-Judge: Claude Opus 4.6 annotated all 1,260 outputs using a structured-output prompt (released in supplementary materials, §4.9). The original protocol envisioned dual independent expert annotation with adjudication; due to clinical workload constraints (acknowledged in §6.2), expert annotation was completed by a single senior clinician on a 51-row priority subset selected to maximize diagnostic value (EGDA edge cases, divergent outputs across arms, and Level 0 queries with ground truth). The expert subset is used to calibrate the LLM-Judge prompt and to estimate inter-rater agreement between the LLM-Judge and clinical judgment; results are reported in §5.2.2. Inter-rater agreement is quantified using percent agreement as the primary statistic and Cohen’s kappa as a secondary statistic, with weighted kappa (linear weights) for the ordinal F4 scale.

To address potential intra-family bias from using Claude Opus 4.6 as the LLM-as-Judge on outputs generated by Claude Sonnet 4, a parallel cross-family annotation pass was performed by Gemini 2.5 Flash (Google) using a byte-identical judge prompt and structured output schema. The two annotation sets are compared in §5.4.2 (Cross-Family Judge Sensitivity Analysis) via Cohen’s kappa per metric. The Claude Opus annotation is reported as the primary scoring layer throughout §5.2–§5.4.1; the Gemini annotation is reported as a sensitivity analysis.

### 4.7 Statistical Analysis

The primary metric is UI rate; secondary metrics include AII, ARR, and factual correctness. Each proportion is reported with a Wilson score 95% confidence interval. Pairwise differences between arms are reported as risk differences (RD) with Newcombe-Wilson hybrid 95% confidence intervals; two-sided p-values for binary outcomes use the chi-square test of independence, with Fisher’s exact test substituted when any cell count is below 5. Because the experimental design is paired (the same case–query pair is run through all three arms), we additionally report McNemar’s exact test (two-sided binomial on the discordant pair counts) for the per-case-per-query binary outcomes for UI Rate and ARR. Per-case binary outcomes for the McNemar tests were derived from the system response text by a deterministic keyword rule (UI = action verb present and refusal language absent on an evidence-insufficient query; ARR = refusal language present on a Level□0 query); the rule and the resulting paired-outcome table are released as supplementary materials so the paired tests can be reproduced from the public final_results.json artifact. Marginal counts under the keyword rule are within close numerical agreement with the LLM-Judge layer for several arm–metric cells (UI Arm A: 84/300 vs. 84/300; ARR Arm A: 151/260 vs. 150/260; ARR Arm B: 160/260 vs. 160/260) and preserve the directional pattern of the LLM-Judge layer for all others; we therefore use the McNemar tests as paired-test confirmation of the chi-square inferences reported on the LLM-Judge layer. AII is the sum of action severity scores (0–3) over evidence-insufficient queries; we report it both as an absolute count and as a per-query rate with an exact Poisson 95% confidence interval, and we test pairwise rate ratios using the conditional binomial test. Inter-rater agreement between the LLM-Judge and the expert clinician is summarized using percent agreement (the primary statistic at this sample size) with Cohen’s kappa reported as a secondary statistic; we note that kappa is unstable when prevalence approaches 0 or 1 [44]. All analyses are two-sided at α = 0.05; given the small number of pre-specified primary contrasts, we report unadjusted p-values and rely on confidence intervals for inference. Analyses were performed in Python 3.11 using SciPy 1.13.

### 4.8 Ablation Studies

To isolate the contribution of each EGDA component, we run two ablated variants against the full proposed system, all on the same 60-case set as the main experiment using the same base model (Claude Sonnet 4) and same evaluation protocol. EGDA-AblV1 removes evidence-level differentiation from the gate, replacing the L0/L1/L2 grading scheme with a binary present/absent check; this isolates the contribution of graded sufficiency. EGDA-AblV2 retains the full L0/L1/L2 grading scheme but removes the comorbidity safety overrides; this isolates the contribution of the comorbidity-related rules and the SAFETY_FLAG output type. Each ablation produces 420 outputs (60 cases × 7 queries × 1 arm). A third ablation conceptually equivalent to “retrieval without an authorization gate” is already represented by the RAG arm (Arm B) of the main experiment. Ablation results are reported in §5.4.3.

### 4.9 Reproducibility

We release query templates, rule libraries, annotation guidelines, the deterministic keyword-scoring script that derives the paired binary outcomes for the McNemar tests (compute_paired_outcomes.py), the resulting paired-outcome table (paired_outcomes_keyword.csv, n = 420), the LLM-as-Judge prompt and structured-output schema, the full annotation files for both judge models (claude_full.jsonl and gemini_full.jsonl, n = 1,260 each), the cross-family agreement script (compute_judge_agreement.py), and analysis scripts (excluding identifiable patient data).

## 5. Results

### 5.1 Experiment Execution

A pilot study of 3 cases (63 API calls, $0.55) confirmed behavioral separation across the three arms. The full main experiment then executed all 60 cases across 3 arms, producing 1,260 system outputs with zero API errors. Main-experiment token consumption was 821,896 input tokens and 570,201 output tokens, at an estimated cost of $11.02. The cross-family judge sensitivity analysis (§5.4.2) added 1,260 Claude Opus 4.6 + 1,260 Gemini 2.5 Flash judge calls at approximately $25 (Anthropic) and $2 (Google). A subsequent Wave 3 expansion produced 1,470 additional system outputs (630 cross-model rerun on Opus 4.6 + 420 EGDA-AblV1 + 420 EGDA-AblV2) and 2,940 additional LLM-Judge annotations (1,470 Claude Opus 4.6 + 1,470 Gemini 2.5 Flash) at a combined cost of approximately $58. Total Anthropic + Google API spend across all stages was approximately $96. All system-output stages used a fixed base model per arm (Claude Sonnet 4 for the main experiment and ablations; Claude Opus 4.6 for the cross-model rerun), the Anthropic API or Google AI Studio API as appropriate, Google Colab as execution platform, fixed system prompts, and a fixed max_tokens setting per stage (2,000 for Sonnet stages, 4,096 for Opus stages). The 3 pilot cases were included in the 60 full-experiment cases.

### 5.2 Primary Results

System outputs were evaluated using an LLM-as-Judge strategy: Claude Opus 4.6 annotated all 1,260 outputs on five dimensions (F1 Correctness, F2 Claim Type, F3 Unjustified Inference, F4 Action Severity, F5 Appropriate Refusal). A senior clinician independently annotated a 51-row priority subset for calibration and inter-rater validation. Table 4 reports the primary results.

#### 5.2.1 Per-Tier Analysis

Table 5 breaks down UI Rate and ARR by evidence completeness tier. EGDA maintained low UI across all tiers: 0.0% at High, 13.0% at Medium, and 7.9% at Low. At the High tier, where complete evidence was available, EGDA achieved 100% ARR on Level 0 queries while allowing definitive answers on Level 2 queries, showing that the authorization gate does not over-refuse when evidence is sufficient.

#### 5.2.2 Expert Calibration

A senior clinician annotated a 51-row priority subset selected to maximize diagnostic value: EGDA edge cases, divergent outputs across arms, and Level 0 queries with ground truth. The LLM-Judge prompt was calibrated against these expert annotations. Table 6 reports inter-rater agreement between the calibrated LLM-Judge and the expert.

On F3, the no-information rate (NIR) baseline—the percent agreement achieved by always predicting the expert majority class “not UI”—is 50/51 = 98.0%, given the F3 expert prevalence of 1/51 (2.0%). The 66.7% agreement observed between LLM-Judge and expert is therefore well below NIR and reflects active disagreement on 17 outputs flagged by the LLM-Judge but not by the expert, rather than chance concordance on a near-deterministic task. The direction of the disagreement is informative: the LLM-Judge consistently over-flags relative to the expert (17 vs. 1), applying an engineering criterion under which a recommendation made on insufficient evidence is unjustified even when clinically reasonable. The LLM-Judge UI Rate estimates reported in §5.2 should therefore be read as upper bounds on a clinically anchored UI rate; the directional comparisons across arms (EGDA vs. RAG vs. LLM-only) hold under either standard, since the same criterion is applied uniformly within each annotation pass. For F1 (Correctness), the 60.8% LLM-Judge–expert agreement reflects a known definitional issue around whether REFUSAL outputs count as correct, rather than measurement noise; F2 (Claim Type, 62.7% agreement, κ = 0.537) and F4 (Action Severity, 68.6% agreement, κ = 0.556) provide moderate kappa support and are the relevant expert-calibration evidence for the claim-classification and severity-weighting components of the framework. The cross-family LLM-Judge analysis (§5.4.2), computed on n = 1,260 rather than n = 51, is not subject to the prevalence-imbalance instability described above and provides the primary evidence of annotation reliability for the headline behavioral safety metrics.

#### 5.2.3 Directional Validation

Four pre-specified directional hypotheses were tested against the primary results in Table 4. Each hypothesis predicts a ranking of arms on a behavioral safety or accuracy measure; each is evaluated using the same statistical framework described in §4.7. Table 7 reports the primary contrast for each hypothesis, the two secondary contrasts, and an evidence-graded verdict.

**Table 7.** Pre-specified directional hypotheses, statistical evidence, and verdicts. RD = risk difference; RR = rate ratio; CI = confidence interval. Tests as defined in §4.7 (chi-square or Fisher’s exact for proportions; conditional binomial on Poisson rates for AII).

| Pre-specified hypothesis | Primary contrast | Secondary contrasts | Verdict |
| --- | --- | --- | --- |
| H1. Unjustified inference rate decreases with stronger constraints (EGDA < RAG < LLM-only). | EGDA vs. LLM-only: RD -40.7% (95% CI -46.9 to -34.0), $p < 0.001$ | EGDA vs. RAG: RD -39.7% (-45.9 to -33.0), $p < 0.001$.<br>RAG vs. LLM-only: RD +1.0% (-7.0 to +8.9), $p = 0.870$. | Partially supported. EGDA significantly lower than both baselines; RAG vs. LLM-only not significantly different (key finding; see §5.3.1). |
| H2. Appropriate refusal rate increases with stronger constraints (EGDA > RAG > LLM-only). | EGDA vs. LLM-only: RD +38.1% (95% CI +31.3 to +44.5), $p < 0.001$ | EGDA vs. RAG: RD +38.1% (+31.3 to +44.5), $p < 0.001$.<br>RAG vs. LLM-only: RD 0.0% (-8.5 to +8.5), $p = 1.000$. | Partially supported. EGDA significantly higher than both baselines; RAG not different from LLM-only. |
| H3. Action inflation index decreases with stronger constraints (EGDA < RAG < LLM-only). | EGDA vs. LLM-only: RR 0.086, $p < 0.001$ (rate per query 0.057 vs. 0.664) | EGDA vs. RAG: RR 0.081, $p < 0.001$.<br>RAG vs. LLM-only: RR 1.061, $p = 0.505$. | Partially supported. EGDA significantly lower than both baselines on AII rate; RAG vs. LLM-only not significantly different. |
| H4. Authorization gate preserves factual correctness (EGDA ≥ RAG > LLM-only). | EGDA vs. RAG: RD +21.9% (95% CI +17.4 to +26.5), $p < 0.001$ | EGDA vs. LLM-only: RD +26.7% (+21.9 to +31.4), $p < 0.001$.<br>RAG vs. LLM-only: RD +4.8% (-1.3 to +10.8), $p = 0.144$ [chi-square]; paired McNemar $p = 0.013$. | Supported with stronger result than predicted: EGDA significantly exceeds RAG by ~22 percentage points, indicating the gate improves rather than trades off accuracy. RAG advantage over LLM-only is detectable only on paired analysis (McNemar $p = 0.013$). |

### 5.3 Subsidiary Findings

#### 5.3.1 RAG Over-Inference Effect

An unexpected finding was that retrieval-augmented generation, despite providing the model with clinical guidelines, failed to systematically reduce unjustified inference relative to the unconstrained baseline. RAG showed no significant improvement on UI Rate (47.7% vs. 48.7%, RD +1.0%, 95% CI −7.0 to +8.9, p = 0.870 [chi-square]; paired McNemar exact p = 0.798 with 32 vs. 29 discordant pairs), no improvement on appropriate refusal (56.9% vs. 56.9%, RD 0.0%, p = 1.000), no improvement on overall refusal output rate (35.2% vs. 36.7%, p = 0.719), and no improvement on action inflation (AII rate 0.705 vs. 0.664, RR 1.061, p = 0.505). Factual correctness improved modestly (74.5% vs. 69.8%; chi-square p = 0.144 not significant, paired McNemar p = 0.013). Per-tier analysis (§5.2.1) shows that RAG produced no significant UI reduction at any tier (High p = 0.064, Medium p = 0.609, Low p = 0.770), and at Medium and Low tiers RAG was numerically worse than LLM-only. Inspection of disagreement cases reveals the mechanism: RAG outputs frequently cited guideline language from the system prompt to justify recommendations for patients whose snapshot data did not support the application of those guidelines. The model treated the guideline corpus as license to recommend rather than as constraint to bound. The pattern was most visible at the Low tier, where data gaps were largest but guideline language still provided a basis for confident-sounding responses. The finding directly supports the core thesis: information supply (retrieval) is not sufficient for inferential governance.

A complementary observation comes from automated keyword scoring (parallel scoring layer alongside LLM-Judge): on the same case-query pairs, RAG triggered the keyword UI flag on 102 outputs versus 84 for LLM-only, with 26 queries where only RAG was flagged versus 8 where only LLM-only was flagged (paired McNemar exact p = 0.003). In other words, retrieval-augmented prompts are more likely than unconstrained outputs to introduce a treatment recommendation that the LLM-only baseline did not produce on the same case-query pair. The keyword-layer reversal is not reproduced by LLM-Judge scoring, which gives RAG more credit for hedged phrasing such as “consider FISH testing” that the deterministic rule classifies as an action recommendation. The two scoring layers nevertheless agree on the qualitative conclusion: RAG without an authorization gate does not improve behavioral safety relative to the unconstrained baseline, and under at least one scoring layer it actively worsens it. RAG provides a knowledge source; EGDA provides a permission rule. The 40-percentage-point UI gap between RAG and EGDA is entirely attributable to the authorization gate, since both arms received identical guideline content.

### 5.4 Robustness and Validation

#### 5.4.1 Cross-Model Robustness Check

To test whether the directional effects are model-dependent, a 30-case subset (10 per tier: BC-H01–H10, BC-M01–M10, BC-L01–L10) was rerun on Claude Opus 4.6 using byte-identical system prompts (lean PROMPT_C, the version used in the main experiment) and an output budget of 4,096 tokens to eliminate truncation. This produced 630 additional outputs (30 cases x 7 queries x 3 arms). Table 8 compares the LLM-Judge–scored results across the two base models on the same 30 cases; both models were scored using Claude Opus 4.6 as the LLM-Judge.

**Table 8.** Cross-model robustness check: Sonnet 4 vs. Opus 4.6 on the same 30-case subset (LLM-Judge scoring; n = 150 insufficient-evidence queries for UI Rate; n = 130 Level-0 queries for ARR).

| **Metric** | **Model** | **LLM-only (A)** | **RAG (B)** | **EGDA (C)** |
| --- | --- | --- | --- | --- |
| UI Rate | Sonnet 4 | 69/150 (46.0%) | 67/150 (44.7%) | 11/150 (7.3%) |
| UI Rate | Opus 4.6 | 116/150 (77.3%) | 113/150 (75.3%) | 27/150 (18.0%) |
| ARR | Sonnet 4 | 103/130 (79.2%) | 79/130 (60.8%) | 125/130 (96.2%) |
| ARR | Opus 4.6 | 33/130 (25.4%) | 33/130 (25.4%) | 78/130 (60.0%) |
| AII | Sonnet 4 | 132 | 154 | 18 |
| AII | Opus 4.6 | 323 | 331 | 118 |

All 12 directional checks (3 metrics x 4 scopes: overall plus High, Medium, Low tiers) were confirmed on Opus 4.6. EGDA maintained the lowest UI Rate (18.0% vs. 77.3% and 75.3%), the highest ARR (79.2% vs. 25.4% and 25.4%), and the lowest AII (118 vs. 323 and 331). The RAG over-inference effect was preserved on Opus: RAG UI (75.3%) was within sampling noise of LLM-only UI (77.3%), and RAG AII (331) was higher than LLM-only AII (323), reinforcing the finding that guideline access without an authorization gate fails to reduce unsafe inference.

Absolute rates differed between models. Opus 4.6 was substantially more prone to over-inference at baseline than Sonnet 4 (UI Rate 75–77% on Arms A/B vs. 46–47% on Sonnet), and EGDA narrowed but did not fully eliminate this gap on Opus (residual UI 18.0% vs. 7.3% on Sonnet; residual ARR 79.2% vs. 96.2% on Sonnet). The cross-model rerun therefore confirms directional consistency (EGDA reduces UI by ≥57 percentage points on both models and raises ARR by ≥34 percentage points on both) while documenting that the absolute level of residual unsafe behavior under EGDA is model-dependent. Compared to a prior 30-case rerun reported in earlier internal drafts that used a NCCN-augmented variant of PROMPT_C and a 2,000-token output budget, the present rerun uses byte-identical main-experiment prompts and a 4,096-token output budget, eliminating prompt mismatch and the 13% truncation rate observed previously on Arm C outputs.

#### 5.4.2 Cross-Family Judge Sensitivity Analysis

To address the concern that the LLM-as-Judge (Claude Opus 4.6) and the system-under-test base model (Claude Sonnet 4) come from the same training family, all 1,260 system outputs were independently re-annotated by Gemini 2.5 Flash (Google) using a byte-identical judge prompt and an identical structured output schema. Gemini 2.5 Flash was selected over Gemini 2.5 Pro after the Pro endpoint produced repeated 503 errors during pilot runs; the Flash model offers comparable structured-output reliability at substantially lower cost. The judge prompt, structured output schema, and full annotation jsonl files for both judge models are released in the project repository (Section 4.9).

Cohen’s kappa was computed per metric between the two annotation sets, treating each judge as an independent rater on the same 1,260 cases. F4 Action Severity used weighted kappa with linear weights given its ordinal scale (0–3); F5 Appropriate Refusal was scored only on the 780 outputs with expected_level = 0, where appropriate refusal is defined. Table 9 reports the results.

**Table 9.** Cross-family inter-rater agreement between Claude Opus 4.6 and Gemini 2.5 Flash on all 1,260 system outputs (or 780 for F5).

| **Metric** | **n** | **% Agreement** | **Cohen’s $\kappa$** | **Interpretation** |
| --- | --- | --- | --- | --- |
| F1 Correctness | 1,260 | 91.7 | 0.723 | Substantial |
| F2 Claim Type | 1,260 | 91.6 | 0.867 | Almost perfect |
| F3 Unjustified Inference | 1,260 | 90.2 | 0.697 | Substantial |
| F4 Action Severity | 1,260 | 91.0 | 0.822<br>(weighted) | Almost perfect |
| F5 Appropriate Refusal | 780 | 93.8 | 0.846 | Almost perfect |

Inter-rater agreement was substantial-to-almost-perfect on all five metrics by Landis and Koch [45] benchmarks. The lowest agreement was on F3 Unjustified Inference ( $\kappa = 0.697$ , percent agreement 90.2%), reflecting boundary cases where Claude Opus more strictly applied the engineering criterion to outputs that smuggled a treatment recommendation into an otherwise hedged or refusing response, while Gemini gave more weight to the surface-level refusal language. The highest agreement was on F2 Claim Type ( $\kappa = 0.867$ ) and F5 Appropriate Refusal ( $\kappa = 0.846$ ), confirming that the qualitative claim taxonomy and the binary refusal decision are stable across model families.

Directional concordance with the primary findings was complete. Both judges produced strict ordering EGDA □ {RAG, LLM-only} on UI Rate and AII, and EGDA □ {RAG, LLM-only} on ARR and F1 Correctness, with all primary contrasts (EGDA vs. each baseline) significant at p < 0.001 in both annotation sets. The principal finding — that an authorization gate produces an order-of-magnitude reduction in unjustified inference relative to information supply alone — therefore holds independent of the LLM-as-Judge model family choice.

A small disagreement on the LLM-only versus RAG contrast was observed. Under Claude Opus annotation, RAG and LLM-only produced statistically indistinguishable UI rates (47.7% vs. 48.7%, p = 0.870). Under Gemini annotation, RAG showed lower UI than LLM-only (22.0% vs. 35.7%, RD −13.7%, p < 0.001). Inspection of disagreement cases reveals that 94 of Gemini’s annotations violated its own rubric by classifying a response as DEFINITIVE or RECOMMENDATION on insufficient evidence while still scoring F3 = 0; the same rubric-application failure occurred only 7 times in the Claude Opus annotation. Re-deriving Gemini’s UI Rate using the rule-consistent classification (any DEFINITIVE or RECOMMENDATION on insufficient evidence counts as F3 = 1) gives Gemini estimates of 35.7% (LLM-only), 40.7% (RAG), and 7.1% (EGDA), which align more closely with the Claude Opus annotation. The qualitative conclusion is robust to either resolution: under no annotation does retrieval reduce UI to the level achieved by the authorization gate, and under all annotations the two non-gated arms differ from EGDA by an order of magnitude.

#### 5.4.3 Ablation Studies

To isolate the contribution of two key EGDA components, we ran two ablated variants of the authorization gate against the full proposed system on the same 60-case set. Each variant differs from the full Arm C system prompt by exactly one component (see §4.8 for the variant definitions); all other elements (extraction stage, no-fabrication rule, JSON output schema, base model Claude Sonnet 4) were held constant. The 420 outputs per variant were scored using the same Claude Opus 4.6 LLM-Judge as the main experiment, with byte-identical judge prompt. Table 10 reports the four primary metrics for each condition.

**Table 10.** Ablation studies: full EGDA vs. two ablated variants on the same 60-case set (LLM-Judge scoring; Sonnet 4 base model). EGDA-AblV1 removes the L0/L1/L2 evidence grading scheme; EGDA-AblV2 removes the comorbidity safety overrides.

| Variant | F1 Correctness (High tier) | UI Rate (insufficient evidence) | ARR (Level-0 queries) | AII (sum) |
| --- | --- | --- | --- | --- |
| EGDA full (Arm C, baseline) | 130/140 (92.9%) | 24/300 (8.0%) | 247/260 (95.0%) | 27 |
| EGDA-AblV1 (no L0/L1/L2 grading) | 119/140 (85.0%) | 82/300 (27.3%) | 200/260 (76.9%) | 77 |
| EGDA-AblV2 (no comorbidity overrides) | 130/140 (92.9%) | 21/300 (7.0%) | 249/260 (95.8%) | 17 |

Removing the comorbidity safety overrides (EGDA-AblV2), in contrast, produced minimal change on UI Rate (8.0% → 7.0%, p = 0.50 by McNemar paired test on identical case-query pairs) and ARR (95.0% → 95.8%). This null result is informative rather than disappointing: the comorbidity overrides serve a distinct function from the evidence grading scheme. Their primary role is to generate SAFETY_FLAG outputs when treatment recommendations are requested without comorbidity data, and to refuse dose-modification queries that lack organ-function data. Because UI/ARR are computed over the broader insufficient-evidence and Level-0 query populations rather than the comorbidity-specific subset, the contribution of these rules is not captured in the primary outcome measures. Their independent value lies in flagging treatment recommendations that would otherwise be issued without consideration of patient-specific contraindications, a behavior captured in the comorbidity_safety_flag field of Arm C outputs but not in the F1–F5 dimensions reported here. The ablation result therefore establishes that the L0/L1/L2 grading scheme alone, not the comorbidity overrides, is responsible for the UI/ARR improvements reported in §5.2.

#### 5.4.4 Cross-Family Agreement on Wave 3 Outputs

The cross-family judge sensitivity analysis reported in §5.4.2 was conducted on the 1,260 main-experiment outputs. To verify that the LLM-Judge reliability holds for the ablation variants and the Opus cross-model rerun, we re-annotated the 1,470 Wave 3 outputs (630 Opus rerun + 420 EGDA-AblV1 + 420 EGDA-AblV2) using both Claude Opus 4.6 and Gemini 2.5 Flash with byte-identical judge prompts. Table 11 reports the agreement statistics.

**Table 11.** Cross-family inter-rater agreement between Claude Opus 4.6 and Gemini 2.5 Flash on the 1,470 Wave 3 outputs (630 cross-model + 840 ablation). Percent agreement and Cohen’sκ [45].

| **Metric** | **n** | **% Agreement** | **Cohen's $\kappa$** | **Interpretation** |
| --- | --- | --- | --- | --- |
| F1 Correctness | 1,470 | 91.4 | 0.628 | Substantial |
| F2 Claim Type | 1,470 | 86.3 | 0.777 | Substantial |
| F3 Unjustified Inference | 1,470 | 86.9 | 0.569 | Moderate |
| F4 Action Severity | 1,470 | 82.8 | 0.674<br>(weighted) | Substantial |
| F5 Appropriate Refusal | 910 | 85.5 | 0.625 | Substantial |

## 6. Discussion and Conclusion

### 6.1 Implications

The primary finding is that an evidence-graded authorization gate reduces unsafe inference by an order of magnitude. EGDA achieved an 8.0% unjustified inference rate compared with 48.7% for the unconstrained baseline and 47.7% for RAG, while simultaneously achieving the highest factual correctness (96.4%). This result addresses a common concern about safety-oriented systems: refusing to answer when evidence is insufficient does not sacrifice accuracy; it improves it, because avoiding confident answers based on missing data eliminates a category of errors.

A secondary finding is that retrieval-augmented generation, without an authorization gate, did not systematically improve inferential safety over the unconstrained baseline. RAG and LLM-only differed by less than two percentage points on UI Rate, by zero percentage points on appropriate refusal, and not significantly on any behavioral safety metric. Providing clinical guidelines without an authorization gate gave the model a vocabulary to rationalize recommendations rather than constraining them; RAG outputs frequently cited guideline language to justify treatment suggestions for patients whose snapshot data did not support the application of those guidelines. This finding directly supports the core thesis: information supply (retrieval) is not sufficient for inferential governance. The 40-percentage-point UI gap between RAG and EGDA is entirely attributable to the authorization gate, since both arms received identical guideline content.

A third finding emerged from the expert calibration process. The clinician annotator identified only 1 of 51 outputs as unjustified inference, while the LLM-Judge identified 17 in the same subset. The gap reflects a definitional difference: the clinician applied a standard where only fabricated data or hallucinated facts count as unjustified, while clinically reasonable advice under incomplete evidence does not. The gap shows that behavioral safety metrics require clinical anchoring, and that the definition of “unjustified inference” is itself a governance decision that should be made explicitly rather than assumed by engineering convention.

A fourth finding comes from the ablation studies in §5.4.3. Replacing the L0/L1/L2 evidence grading scheme with a binary present/absent check (EGDA-AblV1) increased UI Rate from 8.0% to 27.3% (+19.3 percentage points) and reduced ARR from 95.0% to 76.9% (−18.1 percentage points), while removing the comorbidity safety overrides (EGDA-AblV2) produced no detectable change on UI/ARR (8.0% → 7.0%). This component-level decomposition shows that the central mechanism driving EGDA’s behavioral safety improvements is the graded sufficiency stage, not the comorbidity overrides. The comorbidity overrides serve a complementary function (SAFETY_FLAG generation for treatment recommendations missing required clinical context) that is not captured in the F3/F5 dimensions; their independent contribution to safety would require separate evaluation focused on dose-modification and treatment-escalation queries with comorbidity-relevant patient histories. The ablation pattern further reinforces the §5.3.1 argument that information supply alone (Arm B / RAG) cannot substitute for an authorization gate: even an EGDA variant that retains the gate but coarsens its evidence-grading mechanism (AblV1) substantially under-performs the full system.

The framework’s structured output format (evidence set, sufficiency grading, authorization decision, answer, comorbidity safety flag, reasoning) produces an auditable decision trail for each query. Clinicians consulted during framework design noted that this structure could support informed consent processes by explicitly listing missing data and uncertainty, and could provide traceable documentation if clinical decisions are later questioned. While legal implications of AI-assisted decision trails remain unresolved across jurisdictions, the structural capacity for auditability is a prerequisite for any future regulatory framework.

The framework is complementary to the existing guardrails ecosystem described in Section 2.3. Generic guardrails address content safety (toxicity, off-topic responses); evidence-graded authorization addresses inferential safety (whether the evidence justifies the claim). A deployed system could use both.

Beyond the algorithmic contribution, the framework is designed to be deployable within existing clinical decision support infrastructure. The evidence extraction stage (§3.1) consumes a structured snapshot of patient data that maps directly onto a FHIR Bundle of relevant Resources (Condition, Observation, Procedure, MedicationStatement, DiagnosticReport), so the same extraction logic that operates on the registry-derived snapshots in this study can be invoked against a SMART on FHIR app context [33]. The authorization decision and answer can be returned to the EHR as CDS Hooks cards [34]: a definitive answer renders as a recommendation card with the supporting evidence set as the rationale; a Level 0 refusal renders as a “no recommendation possible” card that lists the missing evidence elements rather than a high-priority alert. This distinction matters for alert-fatigue management: the documented 90% override rate of conventional drug-interaction alerts [46] reflects in part a failure to distinguish actionable warnings from low-information notifications. By rendering insufficient-evidence cases as data-request prompts rather than warnings, an EGDA-backed deployment shifts cognitive load onto evidence completion rather than alert dismissal. The structured reasoning trace and authorization rule applied to each query can be persisted as a FHIR Provenance or AuditEvent Resource, providing the audit infrastructure required for the post-deployment monitoring component of the Bates et al. [30] framework. The framework is described here at the integration-design level; we did not implement it within a live EHR in this study, and end-to-end deployment validation in a clinical workflow remains future work.

The framework is built around three components that we expect to transfer across clinical domains, and three that must be reconstructed for each new domain. The first portable component is the architectural separation of evidence extraction, sufficiency grading, and claim-level authorization into three serial stages. The second is the use of discrete, ordered evidence sufficiency levels (here L0–L2) as the input to authorization rules. The third is the principle of binding claim types to minimum required evidence levels through an explicit admissibility condition. The first domain-specific component is the inventory of claim types relevant to the clinical area (here seven, anchored to breast oncology decision points such as staging, molecular subtype, and HER2-targeted therapy candidacy). The second is the evidence types and the criteria that distinguish L0, L1, and L2 for each claim (e.g., what counts as confirmatory pathology versus suggestive imaging). The third is the safety-override rules tied to comorbidities relevant to the planned interventions in that domain. Extending EGDA to, for example, lung cancer, cardiovascular triage, or non-oncologic safety-critical settings would reuse the architecture but require a new claim-type inventory, new sufficiency-level definitions, and new override rules co-developed with domain clinicians; the underlying methodological contribution is the architecture and the admissibility condition, not the specific oncology rule set instantiated here.

### 6.2 Limitations

The experiments used a single base model (Claude Sonnet 4) for the main run and ablation studies, and Claude Opus 4.6 for the cross-model robustness check. The system prompts are model-agnostic and contain no model-specific syntax, but behavioral compliance with structured output instructions may vary across model families. The cross-model robustness check (30 cases, 630 outputs) confirmed all directional effects under the same Claude Opus 4.6 LLM-Judge as the main experiment (§5.4.1), though residual UI Rate under EGDA on Opus (18.0%) was higher than on Sonnet (7.3%), indicating that the absolute level of behavioral safety achieved by the framework is partially model-dependent.

The base model used for the main experiment and both ablation studies (claude-sonnet-4-20250514) was deprecated by Anthropic on 14 April 2026 with end-of-life scheduled for 15 June 2026. We retained Sonnet 4 across the main experiment, the ablation studies, and the LLM-Judge prompt design to maintain controlled comparison and to avoid introducing model-version as a confound when isolating the contribution of EGDA components. Replication on the successor model (Sonnet 4.6 or later) is identified as future work and is one motivation for the cross-model robustness check on Opus 4.6 reported in §5.4.1.

Expert annotation was limited to 51 rows from a single clinician due to clinical workload constraints. This sample size was sufficient to calibrate the LLM-Judge prompt, identify the F3 definitional gap, and establish moderate-to-substantial percent agreement (60.8–90.9% across the five fields), but is too small for stable Cohen’s kappa estimation, particularly for fields with extreme prevalence imbalance (F3 expert prevalence 1/51); for this reason we treat percent agreement as the primary inter-rater statistic and kappa as a secondary statistic with the limitations noted in Table 6. The LLM-Judge served as the primary scoring layer, with moderate kappa support on F2 (Claim Type, κ = 0.537) and F4 (Action Severity, κ = 0.556) against the 51-row expert subset. To address potential intra-family bias from using a same-family LLM-Judge (Claude Opus 4.6 evaluating Claude Sonnet 4 outputs), all 1,260 outputs were independently re-annotated by Gemini 2.5 Flash (Google) using a byte-identical judge prompt and structured output schema; cross-family inter-rater agreement was substantial-to-almost-perfect across all five fields (F1 κ = 0.723, F2 κ = 0.867, F3 κ = 0.697, F4 κ = 0.822 weighted, F5 κ = 0.846; Section 5.4.2). Directional concordance with the primary findings was complete in both annotation sets. Full expert annotation of 1,260 outputs by two independent clinicians, with adjudicated disagreements, would further strengthen the headline estimates beyond what the cross-family LLM-Judge analysis already establishes.

The evaluation relies on cross-sectional clinical snapshots from a hospital cancer registry. These snapshots do not capture the temporal dynamics of real-world clinical workflows, including evolving evidence, retracted findings, or delayed confirmatory results. Validation on longitudinal electronic health record data would improve ecological validity.

The RAG knowledge base used by the RAG and EGDA arms was generated from the model’s training data and verified against publicly available guideline summaries, rather than retrieved from licensed NCCN or AJCC documents (Section 4.2). This was a pragmatic choice driven by licensing constraints, but it may underestimate the performance of a fully provisioned RAG deployment that has access to the official current guideline corpus. The RAG over-inference effect we report should therefore be interpreted as a structural property of guideline access without an authorization gate, rather than a claim that licensed-corpus RAG deployments will necessarily exhibit the same UI Rate. Critically, the RAG and EGDA arms received identical guideline content, so the comparison between them isolates the effect of the authorization gate independent of corpus provenance; the LLM-only versus RAG comparison is the one most affected by this caveat. Replicating the experiment with a licensed guideline corpus would strengthen the LLM-only versus RAG contrast, and we encourage groups with such access to do so.

The framework is developed and evaluated within breast oncology only. Evidence structures, claim types, and comorbidity override rules are domain-specific by design. Extending the framework to other oncologic subspecialties or non-oncologic safety-critical domains requires separate rule development and validation.

The evidence sufficiency levels and claim-action rules are defined by domain knowledge rather than learned from data. This ensures interpretability and auditability but introduces dependence on expert consensus. The current design does not address how rules should be updated as clinical guidelines evolve; a deployed system would need rule versioning and periodic recalibration.

### 6.3 Conclusion

Clinical AI is increasingly deployed in time-pressured oncology settings, where an oncologist may receive a confidently structured LLM output and must decide in minutes whether to act on it. The trust this scenario rests on cannot be built on prediction accuracy alone. What a model is permitted to assert, given the evidence available at the point of decision, is a separate problem from how accurate the model’s parametric knowledge is and from what information it can retrieve. This paper has framed that separate problem as inferential governance and proposed Evidence-Graded Decision Authorization (EGDA) as a concrete instantiation.

The three behavioral safety metrics introduced here (UI Rate, AII, ARR) make the inferential gap measurable at the level of model behavior rather than benchmark accuracy. On 1,260 outputs across 60 breast cancer decision snapshots, EGDA reduced the unjustified inference rate from 48.7% to 8.0%, raised the appropriate refusal rate from 56.9% to 95.0%, and produced the highest factual correctness at 96.4%, showing that an authorization gate improves rather than trades off accuracy. The ablation studies localized the load-bearing mechanism to the L0/L1/L2 sufficiency grading; the architecture, not any single rule set, carries the behavioral safety effect.

The most consequential negative finding is that retrieval-augmented generation without an authorization gate did not reduce any behavioral safety metric relative to the unconstrained baseline (UI Rate p = 0.870; ARR p = 1.000). Current deployment architectures that rely on RAG as their safety mechanism are addressing the information-supply problem, not the inferential-governance problem.

Three lines of future work follow. Architectural portability needs to be tested by reconstructing claim inventories, sufficiency definitions, and override rules for other clinical domains. Ecological validity needs longitudinal EHR data rather than cross-sectional snapshots. Deployment realism needs end-to-end integration through SMART on FHIR and CDS Hooks, with the authorization rule and reasoning trace persisted as Provenance Resources for post-deployment audit.

## Data Availability

The de-identified breast cancer registry data from Changhua Christian Hospital, Taiwan (n=3,999) used in this study are not publicly available due to patient privacy regulations and institutional data sharing policies, but are available from the corresponding author upon reasonable request and subject to institutional approval. Aggregated experimental results, prompt templates, evaluation rubrics, and analysis code are available from the corresponding author upon reasonable request.

## Data availability

This study was approved by the IRB of the participating institution (IRB approval number provided on the title page). The requirement for informed consent was waived given the retrospective nature of the analysis and the use of de-identified records. The clinical registry underlying this work comprises de-identified patient records collected at a tertiary medical center in Taiwan between 2011 and 2019, with all 18 HIPAA-equivalent direct identifiers removed prior to analysis. Due to institutional data-sharing policies and patient privacy regulations applicable to single-center hospital registry data, the underlying registry cannot be made publicly available. The 60 de-identified decision snapshots used in the experimental evaluation, the system prompts for the three experimental arms, the LLM-as-judge prompts, the full experimental code, and the 1,260 (plus Wave 3) system outputs are available from the corresponding author upon reasonable request and execution of a data-use agreement consistent with the IRB protocol.

## Declaration of generative AI and AI-assisted technologies in the writing process

In this work, generative AI was used as the research subject of the experimental evaluation: Claude Sonnet 4 and Claude Opus 4.6 (Anthropic) served as the base models under test, and Claude Opus 4.6 and Gemini 2.5 Flash (Google) served as automated annotators (LLM-as-judge); these uses are described in detail in §4 (Experiments) and are the empirical contribution of this paper. The authors additionally used Claude (Anthropic) for limited language editing during manuscript preparation; the authors reviewed and edited all such content and take full responsibility for the manuscript. No generative AI was used to produce experimental data, statistical analyses, or scientific conclusions; these were performed by the authors using the methodology described in §4.7.

## Ethics approval

The Institutional Review Board of Changhua Christian Hospital, Taiwan gave ethical approval for this work (IRB No. 240616). The requirement for informed consent was waived given the retrospective nature of the analysis and the use of de-identified records.

## CRediT authorship contribution statement

Che Lin: Conceptualization, Investigation, Validation. Jia-Yi Lin: Project administration, Data curation, Visualization, Validation. Yao-San Lin: Conceptualization, Methodology, Software, Validation, Writing – original draft, Writing – review & editing, Supervision.

